# Emotional symptoms, mental fatigue and behavioral adherence after 72 continuous days of strict lockdown during the COVID-19 pandemic in Argentina

**DOI:** 10.1101/2021.04.21.21255866

**Authors:** F Torrente, A Yoris, DM Low, PL Lopez, P Bekinschtein, G Vázquez, F Manes, M Cetkovich

## Abstract

**Background:** An early, total, and prolonged lockdown was adopted in Argentina during the first wave of COVID-19 as the main sanitary strategy to reduce the spread of the virus in the population. The aim of this study was to explore emotional symptoms, mental fatigue, and behavioral adherence associated with the COVID-19 pandemic after an average of 72 days of continuous lockdown in Argentina. Specifically, we intended to know: 1) if the prolongation of the lockdown was associated with elevated emotional symptoms; 2) if the prolonged lockdown affected adherence, a phenomenon called “behavioral fatigue”; and 3) how financial concerns in a developing country affected adherence to the lockdown and emotional status of the population.

**Method:** A survey was designed to evaluate the psychological impact of the pandemic and lockdown. The survey included standardized questionnaires to assess the severity of depressive (PHQ-9) and anxious (GAD-7) symptoms, a questionnaire to evaluate mental fatigue, and several additional instruments to assess other variables of interest: risk perception, lockdown adherence, financial concerns, daily stress, loneliness, intolerance to uncertainty, negative repetitive thinking, and cognitive problems. Three LASSO regression analyses were carried to evaluate the predictive role of the different variables over depression, anxiety, and lockdown adherence

**Results:** The survey was responded by 3617 individuals over the age of 18 (85.2% female) from all the provinces of Argentina. Using the Oxford stringency index, Argentina had one of the most stringent and prolonged lockdowns when the sample was collected with 63 to 77 continuous days with a stringency index of more than 85/100. 45.6% of the sample met the cut-off for depression and 27% for anxiety. Previous mental health treatment, low income, being younger, and being female were associated with higher levels of emotional symptoms. Mental fatigue, cognitive failures, and financial concerns were also associated with emotional and subjective complaints, but not with adherence to the lockdown. In the regression models, mental fatigue, cognitive failures, and loneliness were the most important variables to predict depression, meanwhile intolerance to uncertainty and lockdown difficulty were the most important in the case of anxiety. Perceived threat was the most important variable predicting lockdown adherence.

**Conclusions:** Emotional symptoms persisted and even increased during the extended lockdown, but we found no evidence of behavioral fatigue. Instead, mental fatigue, cognitive difficulties, and financial concerns were expressions of the emotional side of the pandemic and the restrictive measures.

## Introduction

Since the beginning of the COVID-19 pandemic, warnings have been issued about the impact on the mental health of affected populations. The experience of previous epidemics such as 2003 SARS and 2014-2015 Ebola indicated that fear of infection had spread vigorously in populations at risk resulting in elevated levels of health anxiety (1) (2). Also, lockdowns have been shown to provoke negative psychological effects including stress, low mood, irritability, anger and post-traumatic stress disorder in the more severe cases (3). Reports from countries affected earlier by COVID-19 confirmed these presumptions revealing a significant impact of the outbreak on mental health indicators (4) (5) (6) (7). Similarly, in a former study carried out by our group during the first week of the national lockdown in Argentina we found early affective symptoms similar to those reported in other countries (8). Participants evidenced elevated levels of anxiety and depressive symptoms associated with feelings of loneliness, daily stress and repetitive negative thinking as the main explanatory variables. Previous mental health conditions, low income, being younger and female gender were aggravating factors. We interpreted these findings as the consequence of a rapid and forced adaptation to a large and sudden environmental threatening stressor. Increases in health anxiety and behavioral effects of social isolation, reduction in levels of activity, diminished reward, and disrupted routines were invoked as the potential mechanisms to explain the observed emotional outcomes (8). However, as a noticeable difference with previous reports of early psychological impact, our observations occurred when there were few cases and deaths in the country. Strong, generalized and sustained protective measures were adopted in Argentina as the main sanitary strategy to reduce the spread of the virus in the population much in advance in comparison to other nations. By March 20^th^ of 2020, when the national lockdown began in Argentina there were 31 cases and zero deaths in the country (data from COVID-19 Dashboard from John Hopkins University (9)). This decision carried a prominent consequence: a large-scale preventive lockdown extended during several months accompanying the progressive growth of the pandemic. Such a prolonged lockdown created new questions about its effects on the confined population beyond its initial impact. Accordingly, the general aim of this study was to evaluate the emotional, cognitive and behavioral correlates of the extended lockdown in Argentina. As a first question, we intended to explore if the persistence of the stressor was accompanied by a persistence of the emotional symptoms. It has been suggested that the more prolonged the confinement, the more deleterious its consequences (3). However, a longitudinal study in the UK showed that the highest levels of depression and anxiety occurred in the early stages of lockdown but declined soon as most individuals adapted to the circumstances (10). At the same time, other studies suggested that as the pandemic spreads and the contagions rise, the negative emotional correlates also increase (11). Particularly, in Argentina the number of new contagions augmented slowly but steadily with the continuous lockdown in place. Around the time of this study, May, 2020, the first wave of COVID-19 had already hit many northern hemisphere countries at the end of their winter and was just starting to arrive to Latin America that was still benefiting from warmer weather and in cases like Argentina, from early preventative lockdowns. A paradoxical situation was created in which, despite energetic measures, infections continued to increase. Therefore, the two stressors alleged in our previous study, the threat of COVID-19 and the lockdown itself, remained both in force over two months after the beginning of the lockdown.

As a second question, the prolonged lockdown in Argentina created the conditions to explore the controversial issue of behavioral fatigue, whereby individuals begin to ignore regulations they once followed despite ongoing risk, which had even risen in Argentina. Discussions about the concept of behavioral fatigue arose when governments initiated the restrictions in response to the COVID-19 first wave in Europe. Public officials and experts in the UK warned about the possible negative effects of establishing a precipitate lockdown. It was guessed that if the countermeasures were too strict and premature, affected people could develop behavioral fatigue with the undesirable consequence of mining compliance with the sanitary protective measures in general. This idea was heavily criticized by other behavioral scientists who stated that the prediction lacked empirical support (12) (13). However, no study until now reported fatigue measures in the general population during COVID-19 pandemic testing its role as a factor driving non-compliance. In face of the recurrence of new waves of COVID-19 it is important to know how people respond to prolonged restrictive measures.

Finally, a third question concerned the role of socio-economic situation in Argentina, as a developing country struggling against consecutive economic crises, with high prevalence rates of poverty, and many people working in informal positions. The suspension of work and economic activities seem to have created additional stressful life conditions and increasing financial concerns. Consequently, we wanted to evaluate how income and financial concerns interacted with emotional symptoms, mental fatigue and attitudes towards lockdown. Regarding the first question, our hypothesis was that as long as the stressors persist, emotional symptoms would remain stable or increase. In addition, as shown in previous studies, several psychological factors such as feelings of loneliness, intolerance of uncertainty, negative thinking, and daily stress were expected to be associated with the levels of emotional symptoms (8) (14) (15) (16) (17) (18). Concerning the controversial issue of behavioral fatigue, in order to test the predictions of the theory, we adopted as the main hypothesis that mental fatigue would be negatively correlated with compliance with sanitary restrictions, and as the null hypothesis that fatigue would not be correlated with compliance. As a secondary hypothesis, we expected that mental fatigue would be positively correlated with emotional symptoms. With regards to the third question, we expected that financial concerns were associated with emotional symptoms and negatively correlated with lockdown compliance. Finally, we additionally explored which factors explain emotional symptom severity and lockdown adherence.

## Methods

### Participants

This report is based on a sample of 3617 individuals from Argentina over the age of 18. Gender was reduced to three main categories: female, male and other. Education profile was segmented into four categories in line with the national education system (see Table 1 for details). The family’s basic income was asked in monthly Argentine pesos and converted to three categories (low, medium and high income). Due to considerable differences in COVID-19 contagion rate, the sample was divided into two groups “AMBA” (Buenos Aires city and Great Buenos Aires with higher rates) and “non-AMBA” for the rest of the country. As social media platforms were part of the main delivery system, all participants mandatorily gave their informed consent asserting to know their privacy would be protected following the Declaration of Helsinki and national laws. The study was approved by the Ethics Committee of Favaloro Foundation.

**Table 1.** Sociodemographic and COVID-19 related data.

|  |  |  |  |  |
| --- | --- | --- | --- | --- |
| **Age** | **Mean** | **SD** | **Min** | **Max** |
|  | 47.31 | 12.76 | 18 | 95 |
| **Gender (%)** | **Female** | **Male** | **Other** | **NA** |
|  | 85.2 | 14.5 | 0.1 | 0.2 |
| **Education (%)** | **7-12 years** | **12-15 years** | **15-20 years** | **20+years** |
|  | 3.7 | 41.0 | 45.5 | 10.0 |
| **Family income** | **Low** | **Medium** | **High** |  |
|  | 39.3 | 37.4 | 23.4 |  |
| **Perception of COVID-19 threat** | **Very Low** | **Low** | **Medium** | **High** |
| **General (%)** | 12.0 | 21.8 | 24.6 | 41.7 |
| **By ages (%)** |  |  |  |  |
| **18-25** | 29.6 | 27.4 | 25.9 | 17.0 |
| **26-44** | 14.0 | 25.9 | 26.0 | 34.1 |
| **45-64** | 9.6 | 20.2 | 24.4 | 45.8 |
| **65+** | 8.3 | 9.7 | 18.8 | 63.3 |
| **Personal risk of infection** | **Very Low** | **Low** | **Medium** | **High** |
| **General (%)** | 22.5 | 30.6 | 23.8 | 23.0 |
| **By ages (%)** |  |  |  |  |
| **18-25** | 34.1 | 33.3 | 16.3 | 16.3 |
| **26-44** | 23.3 | 31.5 | 23.7 | 21.5 |
| **45-64** | 21.6 | 31.4 | 23.9 | 23.1 |
| **65+** | 19.3 | 22.5 | 27.1 | 30.9 |
| **Adherence to lockdown** | **Yes** | **No** | **Partially** |  |
| **General (%)** | 79.7 | 2.8 | 17.5 |  |
| **By ages (%)** |  |  |  |  |
| **18-25** | 76.3 | 4.4 | 19.3 |  |
| **26-44** | 80.3 | 2.7 | 17.0 |  |
| **45-64** | 78.3 | 2.7 | 19.0 |  |
| **65+** | 85.1 | 3.0 | 11.9 |  |

### Contextual measures: quarantine stringency and mobility

To understand the context the participants of the sample were facing and help explain the mental health measures obtained, we provide data of how Argentina ranks in comparison to other countries in several domains at the time of sample collection. This allows us to provide evidence as to whether the quarantine was particularly stringent or prolonged and whether the death rate or economic situation may be particular to this sample.

#### Stringency

Data was obtained from the Oxford COVID-19 Government Response Tracker (19) which tracks school and workplace closures, public gatherings, travel bans, stay at home exceptions, among other indicators, which should not be interpreted as a rating of the appropriateness or effectiveness of a country’s response, but rather be a more objective comparative measure across countries. Measures were obtained to the midpoint of data collection (05/28/2020) after 70 days of lockdown.

#### Mobility

For a general overview of how mobility decreased during the quarantine, we use data from Google Community Mobility Reports (google.com/covid19/mobility) from users who have turned on the Location History setting. This index is then smoothed to a rolling 7-day average and we grouped subregions into the Metropolitan Area of Buenos Aires and the rest of the country.

### Instruments

A survey was designed to evaluate different variables associated with the psychological impact of the pandemic and lockdown. The survey included two standardized questionnaires to assess the severity of symptoms of the depressive and anxious series, a questionnaire to evaluate mental fatigue, and several additional instruments to assess related variables: **Patient Health Questionnaire-9 (PHQ-9)**. The PHQ-9 is a brief self-report scale composed of nine items based on the DSM-IV criteria for the diagnosis of major depressive episodes. It has been developed to assess the presence and severity of depressive symptoms in primary care and in the community and to establish a tentative diagnosis of a depressive episode. The Argentine version of PHQ-9 (20) had high internal consistency (Cronbach’s alpha□=□0.87) and satisfactory convergent validity with the BDI-II scale [Pearson’s r=□0.88 (p <□0.01)]. The cut-off points established by Urtasun et al. (20) were used to evaluate the possible diagnosis of depression and the ranges of severity in the present study. A score of 8 or more indicated a possible diagnosis of major depression according to DSM-IV. The cut off points for severity ranges were 6–8 for mild cases, 9–14 for moderate, and 15 or more for severe depressive symptoms respectively.

#### Generalized Anxiety Disorder-7 (GAD-7)

The GAD-7 is a brief 7-item self-report questionnaire designed to identify probable cases of generalized anxiety disorder and to assess the severity of symptoms. GAD-7 also proved to have good sensitivity and specificity as a screener for panic, social anxiety, and posttraumatic stress disorder (21). The original version of the questionnaire (22) showed very high internal consistency (Cronbach’s alpha□=□0.92) and satisfactory convergent validity with the Beck Anxiety scale [r=□0.72(p <□0.01)] and with the anxiety subscale of the Symptom Checklist-90 (r = 0.74). The Spanish version for Argentina of the GAD-7 was used in this case (downloaded from: https://www.phqscreeners.com/select-screener). The GAD-7 was utilized in previous studies in Argentina (23) and showed a high internal consistency for the sample of the present study (Cronbach’s alpha□=□0.90). To establish the severity levels of the current sample, the cut-off points were used according to Spitzer et al. (22). Accordingly, scores of 5, 10, and 15 were taken as the cut-off points for mild, moderate and severe anxiety, respectively. A score of 10 or greater was considered as indicative of the presence of a possible anxiety disorder.

#### Mental Fatigue

This term refers to the feeling that people may experience after or during prolonged periods of cognitive activity (24). It could be considered a nonspecific symptomatic dimension reported by patients in many physical diseases and psychiatric conditions, or under workplace stress and other prolonged stressful circumstances. For our study, we employed the five items measuring mental fatigue from the Fatigue Assessment Scale-10 (25). This scale asks respondents how they usually feel in a rating scale from 1 (never) to 5 (always). The original instrument showed an adequate internal consistency of .87. The Cronbach Alpha obtained in our sample for the mental fatigue subscale was .82.

#### COVID-19’s perception and attitudes towards lockdown

The survey included questions created *ad hoc* to evaluate variables related to the pandemic and lockdown. Perception of threat of COVID-19 (severity of its outcomes) and perception of risk of being infected (susceptibility) were explored as two single dimensions in a scale from 0 (“not at all”) to 10 (“extreme”). Attitudes toward lockdown were evaluated considering two self-reported dimensions: compliance with the measure (from 0 “not at all” to 10 “completely compliant”) and subjective difficulty of complying with the lockdown (from 0 “not at all” to 10 “absolutely difficult”).

#### Financial concerns

Present and future financial concerns were explored through two specific single-dimension questions from: 0 “Desperate (can’t afford essentials)” to 10 “Not worried at all”. Present worries correspond to the financial situation at the time of the survey, meanwhile future worries correspond to an estimation of the financial scenario twelve months after. In both cases, a lower rating indicated a more negative evaluation.

#### Daily stress

Impact in daily life was assessed within five domains: work, household chores, physical exercise, leisure, activities with children, and relationship with other adults. For each of the areas, the participants had to rate how difficult it was for them to carry out the daily activities compared to the moments prior to lockdown (from very difficult to very easy). A general index of daily stress was calculated summing up the scores of each of the six domains assessed (with a score of -2 for “very difficult”, -1 for “difficult”, 0 for “neutral”, 1 for “easy”, and 2 for “very easy”). As a result, the index varies from -12 (more negative daily stress scenario) to 12 (more positive daily stress scenario).

#### Feelings of loneliness

Loneliness was measured with the UCLA Loneliness Scale (UCLA-LS (26)) adapted to Argentine Spanish by Sacchi & Richaud de Minzi (27). This 20-item Likert-type scale with 4 options (Never, Rarely, Sometimes Often) asked about self-perception of social connection and negative feelings associated with loneliness as a unidimensional construct. Cronbach’s alpha for our sample was 0.91.

#### Intolerance of Uncertainty - Short Form (IUS-12)

Intolerance of uncertainty (IU) is defined as a “dispositional characteristic that reflects a set of negative beliefs about uncertainty and its implications” (28). People with difficulties tolerating uncertainty have the tendency to believe that uncertainty in itself is distressing, unfair, and should be avoided (29). IU is currently considered a transdiagnostic factor associated not only with worry and generalized anxiety, but also with a more broad array of anxiety and emotional disorders (29). The IUS-12 is a short version of the original 27-item Intolerance of Uncertainty Scale that measures responses to uncertainty, ambiguous situations, and the future (30). The 12 items are rated on a 5-point Likert scale ranging from 1 (not at all characteristic of me) to 5 (entirely characteristic of me). The IUS has been adapted and psychometrically validated in Argentina (31). The Cronbach Alpha obtained in our sample was very high (.92).

#### Negative repetitive thinking

As stated by Ehring and Watkins (32) individuals with emotional disorders usually report excessive and repetitive thinking about their current concerns, problems, past experiences, or worries about the future. In the current study, we explored this dimension by assessing the presence of an increased number of negative thoughts related with past, future, or interpersonal concerns since the beginning of the lockdown. For each of these options there was a categorical (yes/no) answer. Negative repetitive thinking was considered present when at least one of the options was selected.

#### Cognitive failures

This dimension was evaluated with an ad hoc questionnaire that included seven single-dimension questions exploring self-perceived difficulties in attention, planning, memory, and decision-making in the real-life context. Each question was rated in a Likert format with five options from “totally agree” to “totally disagree”. The response was interpreted as positive when participants choose “agree” or “totally agree”. The final index of cognitive failures was the sum of the seven items responses ranging from 0 to 7.

#### Sociodemographic characteristics

Potentially relevant general characteristics such as age, gender, family income, region of the country, and level of education were surveyed. Furthermore, being in current treatment for a previous mental health condition was asked as a proxy for a potential pre-existing disorder.

### Procedures

The survey was distributed through different social media networks (Facebook, Twitter, Instagram, WhatsApp) and through email. The questionnaire was enabled on 21/05/2020 and the recruitment of the present sample was completed in 15 days. The official start of the national lockdown in Argentina was established at 12 am on 03/20/20, so the responses obtained correspond to a period of between 63 and 77 days of restrictions (mean = 72, SD = 4.02).

### Statistical Analysis

Comparisons between groups were made using one-way ANOVA followed by Tukey’s HSD or Tamhane 2 for post hoc comparisons when appropriate. Correlations between measures were carried out by using the Pearson correlation coefficient with Bonferroni correction for multiple comparisons. When analyzing categorical variables, the Pearson chi-square test was used.

LASSO regression was used through the *sklearn* package to develop predictive machine learning models for the two main mental health dependent variables (depression and anxiety) as well as lockdown adherence. We used an 80-20 train-test split. Data was standardized and the L1 penalty coefficient was selected through 10-fold cross-validation on the training set by testing 30 equally spaced values on a log scale from 0.0001 to 1. Then the optimal penalty coefficient was used to train the model and predict the test set. All variables from the pairwise correlation analysis were included in the regression models together with additional independent variables such negative repetitive thinking (ordinal), family income (ordinal), and gender (categorical; converted to two dummy variables for male and female as there were only 9 samples of a different gender identity).

## Results

### Sociodemographic characteristics

The sample size was 3617 individuals. The mean age of participants was 47.31 years (SD = 12.76). Female gender was representated most (85.2 %) and few cases reported a non-binary gender identity or preferred not to answer (0.3 %). There were participants from all country’s provinces. Approximately half of the sample (n = 1765, 48.9%) were from the Metropolitan Area of Buenos Aires (AMBA) where 37% of the country’s inhabitants live. By the first day of the survey, 75.6% of the cases of COVID-19 of Argentina were concentrated in the AMBA. No age differences were found comparing the AMBA (mean = 49.11, SD = 13.45) with the rest of the country (mean = 45.59; SD = 11.81). The sample was well distributed across income levels. Regarding education, even though all ranks are represented, there is a tendency towards over-representing higher education levels. (Table 1)

### Contextual analysis: quarantine stringency and mobility

Argentina ranks 14th out of 184 countries on mean stringency index since the start of data collection (01/01/2020) and 5th on the amount of continuous days with stringent quarantine of 85/100 or more (see Supplementary Material, Table S1 and Figure S1 for the trends of how quarantine stringency increased and oscillated worldwide).

Mobility decreased by 20% to almost 100% during the quarantine while time at home increased to up to 40% as compared to baseline (02/15/2020) (Supplementary Figure S2). The order of largest decreases in mobility where: parks, retail and recreation, transit stations, workplaces, and grocery and pharmacy. Mobility changes were similar between AMBA and the rest of the country. We additionally included measures of infection rate and death rate by COVID-19. At the beginning of the survey there were 9931 cases of COVID-19 in the country and 433 deaths. The final day the cases increased to 20197 and deaths to 608 (9); therefore, the death rate was quite low.

### Perception of COVID-19 and adherence to the lockdown

Regarding COVID-19’s threat perception (Table 1), most participants selected high (41.7 %) and medium (24.6 %) ratings, while low (21.8 %) and very low (12.0%) were the less frequent responses. Segmented by ages, an opposite pattern was found between youngest and oldest participants. Perception of COVID-19 threat was rated as very low by 29.6 % of participants in the 18-25 range, meanwhile only 17 % of them perceived a high threat. In contrast, 63.3 % of older adults (65+) rated the threat as very high, and only 8.3 % estimated a very low threat. Divided by territory, AMBA’s participants showed a higher threat perception than the rest of the country (*F*(1,3615) = 25.16, *p* < .001, eta^2^=.007).

In the case of perceived risk of infection, 30.6 % rated low, 23.8 % medium, 23.0 % high, and 22.5 % very low risk. Segmented by age, results presented a similar trend to perceived threat with older adults perceiving risk of infection more than younger adults, but with smaller differences (Table 1).

Divided by areas, participants from AMBA showed a higher perception of risk of being infected than those from the rest of the country (*F*(1,3615) = 34.64, *p* < .001, eta^2^=.009) where the death rate was higher.

Most participants perceived themselves as compliant with the lockdown (79.7%), meanwhile a minority reported to be partially adherent (17.5%) or non-adherent (2.8%) (Table 1). Partial adherence was higher in the younger group (19.3%) and lower in the older (11.9%) (Table 1). Women were more adherent than men (*F*(1,3606) = 31.27, *p* < .001, eta^2^=.01) and participants from AMBA were more adherent than participants from the rest of the country (*F*(1,3615) = 8.41, *p* = .004, eta^2^=.002). Lockdown adherence was correlated with perceived threat of COVID-19 (r = .249, p < .0001) and perceived risk of infection (r = .171, p < .0001). 41.4% of the total sample experienced the lockdown as highly difficult, meanwhile 24.5% and 34.1% of the sample reported medium and low ratings of perceived difficulty, respectively.

## Question 1: emotional symptoms

### Depression symptoms

PHQ-9’s score analysis revealed that 45.6% of the sample reached the DSM criteria for a major depressive episode. When analyzing the number of symptoms required for the diagnosis according to DSM criteria, 18.3% of participants met the criteria of 5 or more symptoms, including depressed mood or loss of interest. Both indicators support our first hypothesis about the persistence of emotional disturbances during the extended lockdown.

Considering severity ranges, 37.8 % of responders did not show depressive symptoms, followed by 24.5 % with mild, 22.3 % with moderate and 15.4 % with severe symptoms. When divided by age, the younger group (18-25) was the most depressed with 64.4 % of participants reporting moderate and severe scores, followed by 25-44 with 40.1 %, then 45-64 with 35.3 and 65+ with 28.5 % (Table 2).

**Table 2.** Emotional symptoms.

|  | **Total** | **18-25** | **25-44** | **45-64** | **65+** |
| --- | --- | --- | --- | --- | --- |
| **Depression scores (PHQ-9)** |  |  |  |  |  |
| (M/SD) | 8.18 (5.84) | 11.34 (5.73) | 8.60 (5.75) | 7.81 (5.85) | 6.91 (5.59) |
| **Depression severity (PHQ-9)**<br>**(% of the sample)** |  |  |  |  |  |
| None | 37.8 | 14.1 | 34.0 | 41.0 | 48.1 |
| Mild | 24.5 | 21.5 | 25.9 | 23.7 | 23.5 |
| Moderate | 22.3 | 37.0 | 23.8 | 21.3 | 15.2 |
| Severe | 15.4 | 27.4 | 16.3 | 14.0 | 13.3 |
| **Depression diagnosis (PHQ-9)**<br>**(% of the sample)** |  |  |  |  |  |
| Not Depressed | 54.4 | 25.9 | 51.1 | 57.9 | 63.0 |
| Possibly Depressed | 45.6 | 74.1 | 48.9 | 42.1 | 37.0 |
| **Anxiety scores (GAD-7)** |  |  |  |  |  |
| (M/SD) | 7.03 (5.17) | 8.38 (4.94) | 7.52 (5.21) | 6.74 (5.16) | 5.84 (4.83) |
| **Anxiety Severity (GAD-7)**<br>**(% of the sample)** |  |  |  |  |  |
| None | 46.7 | 33.3 | 42.5 | 56.6 | 46.7 |
| Mild | 26.3 | 28.1 | 26.9 | 24.6 | 26.3 |
| Moderate | 16.5 | 27.4 | 18.8 | 11.0 | 16.5 |
| Severe | 10.5 | 11.1 | 11.9 | 7.7 | 10.5 |
| **Anxiety diagnosis (GAD-7)**<br>**(% of the sample)** |  |  |  |  |  |
| Not clinically anxious | 73 | 61.5 | 69.4 | 75.4 | 81.2 |
| Possibly clinical anxiety | 27 | 38.5 | 30.6 | 24.6 | 18.8 |
| **Loneliness Scale**<br>**(% of the sample)** |  |  |  |  |  |
| Low | 66.3 | 43.0 | 66.4 | 68.0 | 67.1 |
| Medium | 17.1 | 28.1 | 18.1 | 16.0 | 14.4 |
| High | 10.4 | 20.0 | 10.3 | 9.6 | 10.2 |
| Extreme | 6.2 | 8.9 | 5.2 | 6.3 | 8.3 |

Between groups comparison (one-way ANOVA) showed that participants in current mental health treatment had significantly higher levels of depression (*F*(1,3615) = 103.56, *p* < .001, eta^2^ =.028). However, participants without current treatment also exhibited elevated rates of depressive symptomatology, since 42.2 % of them scored above the cut-off for a possible diagnosis of depression and 34.3 % rated moderate and severe levels of depressive symptomatology.

There were also differences in depression scores between age groups (*F* (3,3613) = 24.29, *p* < .001, eta^2^ = .025). Post-hoc comparisons showed differences between all groups (18-25 > 26-45 > 46-64 > 65+). As well, females were significantly more depressed than males (*F*(2,3614) = 14.13, *p* < .001, eta^2^ = .008). Finally, significant differences in depression scores according to income were found (*F* (1,3615) = 50.57, *p* < .001, eta^2^ = .013). Post-hoc comparisons revealed differences between the three groups (low income > medium > high; *p* < .001). Depression showed no differences between AMBA and the rest of the country (*p* < .14).

### Anxiety severity

GAD-7’s global ratings showed that 46.7 % of participants scored minimal or no anxiety symptoms, followed by 42.8 % with mild and moderate symptoms and 10.5 % with severe symptoms (Table 2). A possible DSM diagnosis for an anxiety disorder (>10) was found in 27 % of the total sample. Thus, ratings of anxiety also support our hypothesis concerning the persistence of emotional symptomatology during the extended lockdown. Divided in age subgroups, the 18-25 and 25-44 groups displayed the higher rates of anxiety (38.5 % and 39.7 % with moderate or severe symptoms, respectively), followed by 45-64 (24.6 %) and 65+ (18.8 %). As in the case of depression, being on current treatment for a mental health condition (*F*(1,3615) = 144.79, *p* < .001, eta^2^ = .039) and being a woman (*F*(2,3614) = 11.42, *p* < .001, eta^2^ = .006) were associated with higher levels of anxiety. Comparison between income groups revealed significant differences in anxious scores (*F*(2,3614) = 16.76, *p* < .001, eta^2^ = .009). Post-hoc comparisons revealed differences between the three groups (low > medium = high; *p* < .001). Anxiety showed no group differences between AMBA and the rest of the country (p = .52).

### Psychological correlates of emotional symptoms

38.1% of the sample endorsed a negative global daily life stress index. Doing physical exercise (50.1 % of participants rated “difficult” or “very difficult”; note parks were closed and jogging was banned in many parts of the country), engaging in leisure activities (44.7 %), and accomplishing work duties (41.7 %) were the more difficult experiences. There were no differences in daily life stress between AMBA and the rest of the country’s (p = .42). Daily life stress was correlated with depression (r = -.306, p < .0001) and anxiety scores (r = -.292, p < .0001) (Table 3).

**Table 3.** Correlations.

|  | 1 | 2 | 3 | 4 | 5 | 6 | 7 | 8 | 9 | 10 | 11 | 12 | 13 |
| --- | --- | --- | --- | --- | --- | --- | --- | --- | --- | --- | --- | --- | --- |
| 1. Depression (PHQ-9) | 1 |  |  |  |  |  |  |  |  |  |  |  |  |
| 2. Anxiety (GAD-7) | .743* | 1 |  |  |  |  |  |  |  |  |  |  |  |
| 3. Mental fatigue | .644* | .525* | 1 |  |  |  |  |  |  |  |  |  |  |
| 4. Cognitive failures | .549* | .491* | .495* | 1 |  |  |  |  |  |  |  |  |  |
| 5. Intolerance of Uncertainty | .436* | .478* | .384* | .326* | 1 |  |  |  |  |  |  |  |  |
| 6. Loneliness Scale | .520* | .481* | .467* | .446* | .341* | 1 |  |  |  |  |  |  |  |
| 7. Daily stress | -.306* | -.292* | -.247* | -.192* | -.177* | -.239* | 1 |  |  |  |  |  |  |
| 8. Age | -.124* | -.107* | -.149* | -.008 | -.063* | -.034 | .050 | 1 |  |  |  |  |  |
| 9. Perceived Threat | .023 | .040 | .005 | .054 | .053* | .039 | 0.01 | .228* | 1 |  |  |  |  |
| 10. Perceived Risk | .068* | .092* | .044 | .068* | .104* | .056 | -.040 | .088* | .561* | 1 |  |  |  |
| 11. Lockdown Adherence | -.060* | -.049* | -.059* | -.060* | .028 | -.042* | .043 | .038 | .249* | .171* | 1 |  |  |
| 12. Lockdown difficulty | .330* | .356* | .224* | .246* | .153* | .251* | -.265* | -.025 | -.023 | .028 | -.051 | 1 |  |
| 13. Financial concerns (present) | -.237* | -.228* | -.155* | -.149* | -.084* | -.168* | .022 | .048 | .012 | .003 | .101 | -.129* | 1 |
| 14. Financial concerns (future) | -.262* | -.262* | -.203* | -.185* | -.126* | -.178* | .081* | .072* | -.001 | -.036 | .077 | -.158* | .732* |

\*. $p < .0001$ and are significant after Bonferroni correction for multiple comparisons (corrected $\alpha = 0.0003$ )

Regarding subjective loneliness, 66.3 % of the sample scored low.17.1 % medium and 16.6 % high ratings at the UCLA loneliness scale. More intense feelings of loneliness appeared in the group aged 18 to 25 years (28.9% in high and extreme grades) than in the groups of 26 to 64 years (15.5%) and 65+ years (18.5%). AMBA showed significant lower levels of loneliness (*F*(1,3615) = 4.829, *p* = .028, eta^2^ = .001) than the rest of the country. Loneliness was positively correlated with scores of depression (r = .52, p < .01) and anxiety (r = .48, p < .01) (Table 3).

73.6% of the total sample expressed at least one kind of negative repetitive thinking (NRT) during the period of lockdown, with higher rates for younger participants (87.4 %). The group with NRT had significantly higher depressive (*F*(1,3615) = 481.05, *p* < .001, eta^2^ = .117) and anxiety scores (*F*(1,3615) = 521.77, *p* < .001, eta^2^ = .126) than the group without NRT.

Finally, intolerance of uncertainty was positively associated with both depression and anxiety (r = .436, p < .0001 and r = .478, p < .0001, respectively).

## Question 2: Mental fatigue and cognitive failures

Contrary to the main hypothesis of behavioral fatigue, mental fatigue was not correlated with adherence to lockdown (r = -.059, p < .0001; Table 3). However, mental fatigue was positively correlated with depression and (r = .644, p < .0001) and anxiety (r = .525, p < .0001) supporting our secondary hypothesis. Even if mental fatigue was not associated with adherence, it was positively correlated with lockdown difficulty (r = .224, p <.0001). There was no difference in mental fatigue between AMBA and non-AMBA participants (p = .30).

Cognitive failures were positively correlated with depression (r = .549, p < .0001), anxiety (r = .491, p < .0001), and mental fatigue (r = .495, p < .0001). Also, 62.48% of the sample experienced failures in three or more cognitive areas.

## Question 3: Financial concerns

As expected, both present and future financial concerns were associated with depression, anxiety, mental fatigue, cognitive failures, and loneliness (Table 3). In contrast, financial concerns were not correlated with adherence to the lockdown. There were differences in present and future worries between the three income groups (F(2,3614) = 180.32, p < .001, eta^2^ = .091; low > medium > high).

### Predictive models of depression, anxiety, and lockdown adherence

Three LASSO regression analyses were carried to evaluate the predictive role of the different variables over depression, anxiety, and lockdown adherence (Table 4).

**Table 4.**
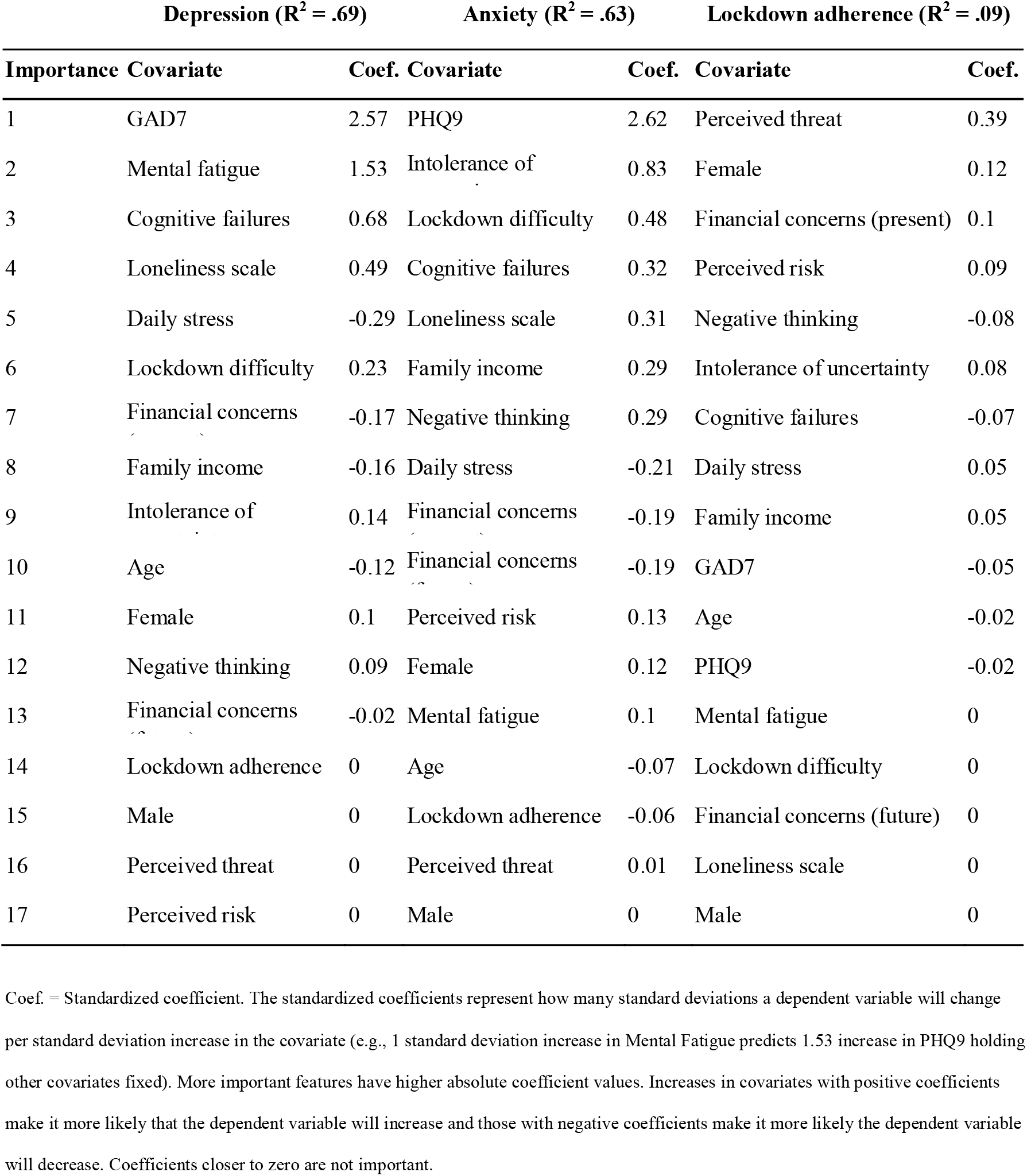
Variable importance when predicting outcome variables using LASSO regression.

The covariates explained 69% of the variance when predicting depression (PHQ9 total scores) and the most important variables, in order, were anxiety (GAD7 total scores), mental fatigue, cognitive failures, loneliness scale, family income, daily stress, and lockdown difficulty. When predicting anxiety (GAD7), the covariates explained 63% of the variance and the most important variables were depression (PHQ9), intolerance to uncertainty, lockdown difficulty, cognitive failures, loneliness scale, family income, and negative thinking. Finally, the covariates explained only 9% of the variance of lockdown adherence predictions and the most important variable was perceived threat.

## Discussion

The aim of this study was to evaluate the emotional and cognitive correlates of the COVID-19 pandemic alongside a lockdown extended between 63 and 79 days in Argentina. Regarding the first question related to the incidence of emotional symptoms, as hypothesized, to the extent that things went wrong and the spread of the virus increased steadily despite the established lockdown –as it had arrived later in Latin America–, the emotional states were consistent with the sustained negative scenario. Ratings of depression and anxiety were higher than those observed in a previous report two months before (45.2 versus 33.7% for depression, and 27 versus 23.2% for anxiety (8)). In agreement with the former report, previous mental health treatment, low income, younger age, and female were associated with higher levels of depression and anxiety. Since the sample is not the same as the previous report, we cannot discard that the increment may be due to sampling differences or other potential confounding variables. However, the observed rates of symptoms are high enough to assert that the psychological impact of the pandemic remained during the lockdown and that there was no habituation or adaptation to the situation. We interpret these findings as an indication that the emotional state of the population accompanies the trajectory of the pandemic and the rate of infection.

Regarding the second question, the issue of behavioral fatigue, we observed that mental fatigue was highly correlated with emotional symptoms and was the second most important variable for predicting depression after anxiety symptoms, and that after an average of 72 days of confinement mental fatigue was associated with perceiving the lockdown as a difficult experience. Notwithstanding, in contradiction with the hypothesis about the behavioral effects of mental fatigue, most people were highly compliant with the imposed restrictions. The main effect of mental fatigue was manifest at the emotional level, instead of the behavioral level, at least under the range of 63 and 77 days covered by the present study. According to the present findings, feeling mentally fatigued is not the same as being behaviorally fatigued.

Related to our third question about financial concerns, while it may be plausible that worrying about the economy would be associated with relaxing lockdown adherence, we did not find this effect: financial concerns were not associated with lockdown adherence. financial concerns correlated with moderately correlated emotional symptoms, and were not important variables when taking into account other covariates in the regression models. However, family income appears to contribute to the prediction of anxiety, albeit slightly. Together these findings indicate that prolonged restrictions have an emotional cost, but people may feel compelled to maintain the safety behaviors for different reasons despite fatigue and financial concerns. In our study, only perceived threat of the COVID-19 and perceived risk of infection correlated with lockdown adherence. This fact, alongside the significant levels of anxiety registered in the population, may imply that threat appraisal is the main relevant driver for people to adopt protective measures. High anxiety scores may suggest that people were still worried about the threat of COVID-19 as the number of cases continued to increase at the time of the survey, and this fear could have reinforced lockdown compliance. The perceived threat (severity of the disease) and the risk of infection (susceptibility) could be understood as the cognitive components of the appraisal, while anxiety reflects its emotional correlate (33). Consistent with these findings, prior to the COVID-19 outbreak, a review by (34) on the determinants of protective behaviors during pandemics found that higher levels of perceived susceptibility and perceived severity of diseases, as well as higher state anxiety levels are associated with preventive behaviors. Even though mental fatigue did not appear to be related with lockdown adherence, it was found to be the second most important factor predicting depression in the regression model. In the context of the pandemic, mental fatigue can result from prolonged cognitive effort to maintain safety behaviors and from sustained inhibitory control to restrict habitual behaviors, such as close physical contact. Also, the hypervigilance associated with anxiety and fueled by the incessant flow of threatening news can lead to mental fatigue. Cognitive failures, another factor associated with depression, may be linked to mental fatigue, since research shows that fatigued subjects have difficulties in focusing their attention, planning, and adaptively changing strategies in the face of negative outcomes (24).In addition, as we observed in our previous report (8), the feeling of loneliness was another important factor related to depression, possibly magnified by prolonged social restrictions for specific groups at risk (35). In the case of anxiety, the regression model supported the significant role that intolerance of uncertainty plays in generalized anxiety and worry according to cognitive-behavioral models (29) (36) (28).

Interestingly, there were no differences in emotional distress and mental fatigue between AMBA and the rest of the country, even if the former region was much more affected by COVID-19. A possible explanation entails the exposure to national news and social media that may have activated negative emotions in advance (37).

Facing the new waves of COVID-19, this study has several potential practical implications. First, there is no conclusive evidence in our study for supporting the phenomenon of behavioral fatigue after an average of 72 days of stringent lockdown. Despite the high levels of fatigue and emotional symptoms, most people strongly adhered to the established lockdown. A plausible interpretation of this fact is that the perceived threat and perceived risk of infection were the main drivers of the high adherence to the lockdown amid its negative emotional consequences. Therefore, a sustained societal awareness of the persistence of the perils of the pandemic could constitute a relevant factor to maintain protective behaviors or to instantiate new restrictive measures. At the same time, a strong fear-promoting communication focusing on negative outcomes and threats may worsen the emotional status of the population even more. As noted by Bish and Michie (34), communications designed to highlight perceptions of risk should also be combined with advice as to how the threat can be alleviated. Furthermore, since mental fatigue may be indicating that people have a negative subjective experience of the lockdown despite being adherent, it may be helpful to communicate clear examples and simple figures of how effort invested leads to positive results.

A second practical implication is that public health strategies should be considered to ensure an adequate coverage of the wide range of unprecedented mental health necessities created by the pandemic. These include warranting the provision of mental health treatment to individuals with pre-existing conditions, identifying and assisting vulnerable groups, allowing a rapid detection and access to care for new emerging cases, and finally promoting preventive behaviors oriented to the wellness of the general population.

The present study has several limitations. First, the survey was disseminated incidentally. Nevertheless, all the country’s provinces were sampled. Also, as the survey was disseminated through social media networks and e-mails, it is possible that a bias occurred towards participants with higher education and income levels. Second, as noted previously, self-report methods may overestimate the rate of psychiatric disorders in comparison with the more reliable gold standard of diagnostic interviews. A recent meta-analysis about the use of the PHQ-9 for the screening of major depressive episodes in primary care found that approximately half of patients with positive screens could be false positives (38). It is important to prudentially consider the present results and to avoid jumping into clinical conclusions. Complementary and more precise procedures should be adopted to confirm or reject any assumption of diagnosis. Third, our sample was unbalanced in gender, with females being overrepresented over other options. Female gender is associated with increased rates of anxiety and depression in epidemiological studies, so sampling bias may have inflated the figures of emotional symptoms of our study. Fourth, due to the observational nature of the study, it is not possible to disentangle the effects produced by the pandemic itself from the impact of the lockdown. Our analyses are not intended to estimate the causal effect between variables nor be an exhaustive analysis of the context (e.g., quarantine stringency, mobility, death rate) on mental health symptoms. The contextual analysis provides quantitative evidence that at the time of the study, Argentina had one of the most continuously stringent lockdowns with low mobility but also low death rate. A more exhaustive analysis could include measures of unemployment, Gini index for income inequality, intensive care unit occupancy, stimulus packages that could help explain symptoms; however, some of these measures are not yet available and are not the focus of the analysis.

In conclusion, emotional distress persisted during the extended lockdown as evidenced by the high levels of depression and anxiety symptoms, but we found no evidence of behavioral fatigue given lockdown adherence remained high. Rather, mental fatigue, cognitive failures, intolerance of uncertainty, loneliness, and financial concerns may be considered as components of the emotional impact of the pandemic.

## Data Availability

Data will be openly available after peer reviewed publishing.

## Declaration of Interest

None.

## Funding Statement

This work was supported by the Interamerican Development Bank (RG-T3106) and the INECO Foundation.

## Author Contribution

F.T. and A.Y. conceived the design, performed data analysis, and drafted the manuscript. D.M.L. contributed to data analysis and writing. All authors provided feedback on design, analyses and reviewed the manuscript.

## Data availability

Code and data will be openly available after peer-reviewed publishing.

## Notes

### Competing Interest Statement

The authors have declared no competing interest.

### Author Declarations

The study was approved by the Ethics Committee of Favaloro Foundation.

