## Supplemental Material for "Emotional symptoms, mental fatigue and behavioral adherence after 72 continuous days of strict lockdown during the COVID-19 pandemic in Argentina"

### 1. Contextual analysis

#### 1.1. Stringency

**Table S1. Countries with most prolonged strict quarantines (continuous days with a quarantine stringency index above 85).**

| Rank | Country | Days |
|---|---|---|
| 1 | Kosovo | 75 |
| 2 | Honduras | 74 |
| 2 | Peru | 74 |
| 2 | Puerto Rico | 74 |
| 3 | Ecuador | 73 |
| 4 | Philippines | 72 |
| 5 | Argentina | 70 |
| 5 | Dominican Republic | 70 |
| 5 | Morocco | 70 |
| 6 | Bolivia | 69 |
| 6 | El Salvador | 69 |
| 7 | Guatemala | 68 |
| 8 | Saudi Arabia | 67 |
| 9 | Nepal | 66 |
| 9 | Libya | 66 |
| 9 | Bahamas | 66 |
| 10 | Paraguay | 65 |
| 10 | Palestine | 65 |
| 10 | Ukraine | 65 |
| 11 | Kenya | 63 |

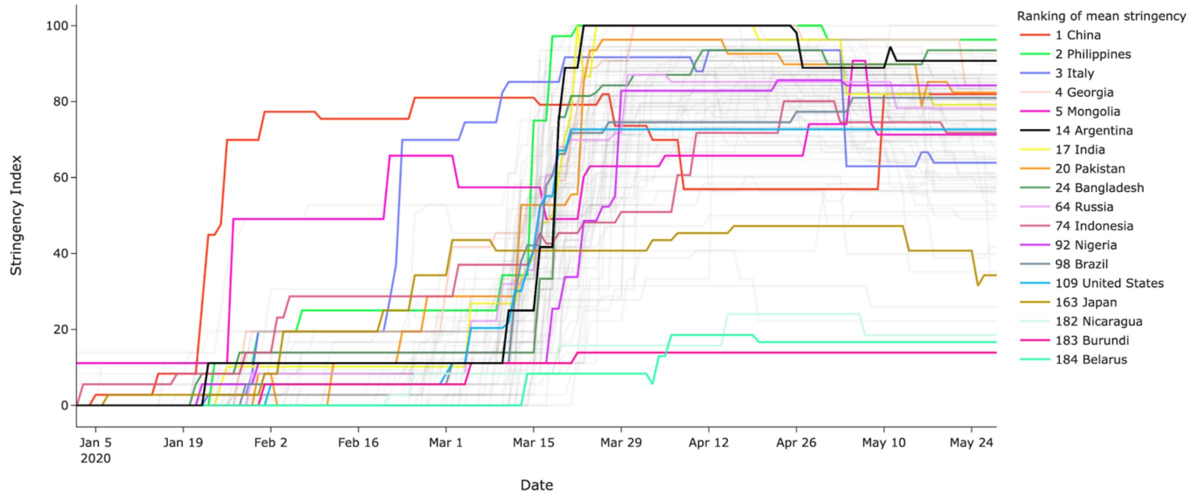


**Figure S1. Stringency Index over time**. Shown are top five ranked countries by mean stringency index, Argentina, ten largest countries in population and bottom three ranked countries. Note Argentina did not lower its stringency index below 85/100 once the lockdown began, placing it in the top ten countries with longest stringent quarantine (see Supplementary Table S1).

### 1.2. Mobility

Mobility trends are measured across six broad categories: (1) Residential: places of residence. (2) Grocery & Pharmacy stores: places like grocery markets, food warehouses, farmers markets, specialty food shops, drug stores, and pharmacies. (3) Workplaces: places of work. (4) Parks: places like local parks, national parks, public beaches, marinas, dog parks, plazas, and public gardens. (5) Transit stations: places like public transport hubs such as subway, bus, and train stations. (6) Retail & Recreation: places like restaurants, cafes, shopping centers, theme parks, museums, libraries, and movie theaters.

See Supplementary Figure S1 to observe how mobility decreased by 20% to almost 100% during the quarantine while time at home increased to up to 40% as compared to baseline

(02/15/2020). The order of largest decreases where: parks, retail and recreation, transit stations, workplaces, and grocery and pharmacy.

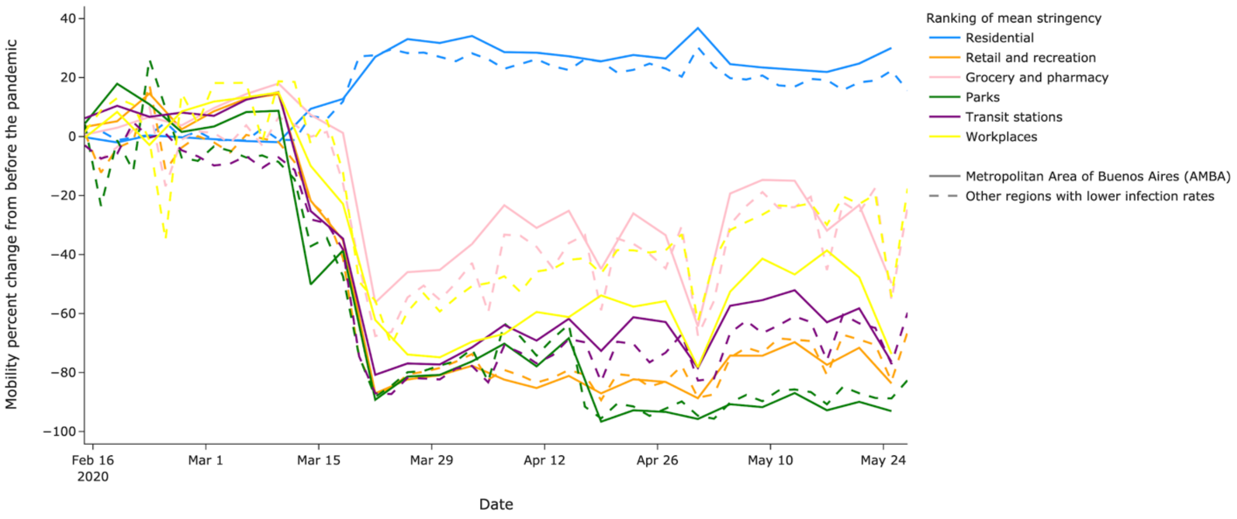


**Figure S2. Decrease in mobility during the quarantine in Argentina and increase in the time spent at home.**
